# Ocular manifestations and clinical characteristics of 534 cases of COVID-19 in China: A cross-sectional study

**DOI:** 10.1101/2020.03.12.20034678

**Authors:** Liwen Chen, Chaohua Deng, Xuhui Chen, Xian Zhang, Bo Chen, Huimin Yu, Yuanjun Qin, Ke Xiao, Hong Zhang, Xufang Sun

## Abstract

**Objective:** The novel coronavirus disease (COVID-19) was first reported in Wuhan, China in December 2019 and is now pandemic all over the world. Previous study has reported several COVID-19 cases with conjunctivitis. However, the complete profiling of COVID-19 related ocular symptoms and diseases are still missing. We aim to investigate the ocular manifestations and clinical characteristics of COVID-19 patients.

**Methods:** A total of five hundred and thirty-four patients were recruited at Mobile Cabin Hospital and Tongji Hospital. We collected information on demographic characteristics, exposure history, ocular symptoms, systemic concomitant symptoms, eye drop medication, eye protections, radiologic findings, and SARS-CoV-2 detection in nasopharyngeal swabs by RT-PCR from questionnaires and electronic medical records.

**Results:** The median age of patients was 40 and 50 years at Mobile Cabin Hospital and Tongji Hospital, respectively. Of 534 COVID-19 patients, 25 patients (4.68%) presented with conjunctival congestion and 3 patients had conjunctival congestion as the initial symptom. The average duration of conjunctival congestion was 4.9 ± 2.6 days (mean [SD]), ranging from 2 to 10 days. Dry eye (112, 20.97%), blurred vision (68, 12.73%), and foreign body sensation (63, 11.80%) ranked as the top three COVID-19 related ocular symptoms. Notably, a total of 332 COVID-19 patients (62%) had a hand-eye contact history. We also found that some COVID-19 patients had a history of eye disease, including conjunctivitis (33, 6.18%), dry eye (24, 4.49%), keratitis (14, 2.62%), cataract (9, 1.69%), and diabetic retinopathy (5, 0.94%). In consistent with previous studies, the most common clinical symptoms were fever, cough, and fatigue. Patients, 60.5% in Mobile Cabin Hospital and 67.5% in Tongji Hospital, respectively were confirmed with positive SARS-CoV-2 detection.

**Conclusions:** Conjunctival congestion was one of the COVID-19 related ocular symptoms, which may have clinical diagnostic significance. It is essential to provide eye-care equipment and strengthen education on eye protection, as dirty hand-eye contact might be a high risk factor of COVID-19. Further detailed and comprehensive ophthalmological guidance is needed for COVID-19 control.

## Introduction

The ongoing outbreak of the novel coronavirus disease (COVID-19) has been declared by WHO as a global public health emergency. COVID-19 was first reported in Wuhan, China, in December, 2019, followed by an outbreak across Hubei Province, other parts of China, and now all over the world, particularly in South Korea, Iran, Italy, and Japan. A study in the by Zhong Nanshan and colleagues^1^ reported nine cases with conjunctival congestion among the 1,099 cases enrolled. However, other ocular manifestations of COVID-19 are unknown, such as increased conjunctival secretion, epiphora, and diminution of vision. Several infected cases^2, 3^ presented firstly with conjunctivitis before the onset of pneumonia, implying that the ocular route might be the potential transmission route of SARS-CoV-2 virus under certain conditions. A recent study revealed that SARS-CoV-2 also uses the cell entry receptor, angiotensin converting enzyme 2 (ACE2),^4, 5^ similar to SARS-CoV. Moreover, the expression and activity of ACE2 can be detected in the ocular surface, including the cornea and conjunctiva,^6^ which provides transocular entry potential for SARS-CoV-2.

Several urgent questions need to be addressed, including 1) What are the detailed profiles of COVID-19–related ocular symptoms and diseases? 2) Do COVID-19 patients with ocular symptoms progress differently from patients without ocular symptoms? and 3) Can COVID-19 spread through the ocular route or present as the primary infected site? To answer these questions, it is essential to perform ocular screening among patients with COVID-19. To our knowledge, comprehensive ophthalmological data on COVID-19 is still missing.

Hence, the present cross-sectional study was designed to describe the ophthalmic characteristics of COVID-19 patients in Wuhan, aiming to get a complete ocular screening of COVID-19, which may provide clinical clues for the diagnosis and treatment of the disease and a theoretical basis for appropriate protection guidelines in the population.

## Methods

### Patients

Five hundred and thirty-four patients diagnosed with novel coronavirus pneumonia (COVID-19) were recruited from February 1 to March 1, 2020, at Mobile Cabin Hospital of Optical Valley and Tongji Hospital of Huazhong University of Science and Technology in Wuhan, China. The ethics committee of Tongji Hospital and the China Ethics Committee for Registering Clinical Trials (ChiCTR2000030489) approved the study. Written informed consent was obtained from the patients involved. An electronic questionnaire were designed to collect demographic, clinical, and ophthalmic data from patients. If data were missing from the questionnaire or clarification was needed, we communicated directly with the patient by telephone after obtaining informed consent. Diagnosis and classifications of COVID-19 were made according to the novel coronavirus infection pneumonia diagnosis and treatment guideline, 7th edition, published by the National Health Commission of China.^7^

### Data collection

For the inpatients at Tongji Hospital, demographic, epidemiological, clinical, laboratory, radiologic, and outcome data were obtained from patients’ electronic medical records. Data about ocular signs and symptoms and the use of eye drops were obtained by ophthalmologists via telephone. For the patients at Mobile Cabin Hospital of Optical Valley, all the data mentioned above were collected via face-to-face survey and an electronic questionnaire completed by patients on a smartphone. Two researchers independently reviewed the data collection forms to double-check the data collected. For uncertain epidemiological and symptom data, the researchers directly communicated with patients or their families via telehone to supplement the data.

### Sample collection

Nasopharyngeal swabs were collected from all patients by a trained healthcare worker using protective equipment. The swab samples were maintained in a viral-transport medium stored between 2° C and 8° C and were analyzed by real-time RT-PCR.

### Nucleic acid detection of SARS-CoV-2

All samples were analyzed by conventional qualitative reverse transcription-polymerase chain reaction (RT-PCR). RNA was extracted from the clinical samples using a viral RNA kit. A 25–μL reaction containing 5 μL of RNA, 12.5 μL of 2× reaction buffer provided with the one-step RT-PCR system with Platinum Taq Polymerase (Invitrogen, Darmstadt, Germany; containing 0.4 mM of each deoxyribont triphosphates (dNTP) and 3.2 mM magnesium sulphate), 1 μL of reverse transcriptase/Taq mixture from the kit, 0.4 μL of a 50–mM magnesium sulphate solution (Invitrogen), and 1 μg of nonacetylated bovine serum albumin. The SARS-CoV-2 specific primers are as follows^8^: forward primer 5′-ACTTCTTTTTCTTGCTTTCGTGGT-3′; reverse primer 5′-GCAGCAGTACGCACACAATC-3′; and the probe 5′CY5-CTAGTTACACTAGCCATCCTTACTGC-3′BHQ1. These primers were used to detect the presence of SARS-CoV-2 RNA. All oligonucleotides were synthesized and provided by Tib-Molbiol (Berlin, Germany). The RT-PCR reaction condition was developed by the Clinical Laboratory of Tongji Hospital. Thermal cycling was performed at 55° C for 10 min for reverse transcription, followed by 95° C for 3 min and then 45 cycles of 95° C for 15 s and 58° C for 30 s.

### Statistical analysis

Continuous variables were expressed as medians and ranges. Categorical variables were summarized as counts and percentages. No imputation was made for missing data. The statistics were descriptive only because the enrolled patients in our study were not derived by random selection. All the analyses were performed using Empower (R) (www.empowerstats.com, X&Ysolutions, Inc. Boston MA) and R (http://www.R-project.org).

## Results

### Demographics and baseline characteristics of COVID-19 patients in Mobile Cabin Hospital and Tongji Hospital Hospital

A total of 263 COVID-19 patients in Mobile Cabin Hospital and 271 COVID-19 patients in Tongji Hospital were enrolled in the study. The demographic data, exposure history, and past medical history are summarized in Table 1.

**Table 1.** Demographics and baseline characteristics of COVID-19 patients in Mobile Cabin Hospital and Hospital.

| **Demographics and baseline characteristics** | **Mobile Cabin Hospital**<br>**(N=263)** | **Hospital**<br>**(N=271)** |
| --- | --- | --- |
| Median age (range) — yr | 40 (16-68) | 50 (19-65) |
| Age group — no./total no. (%) |  |  |
| <15yr | 0/263 | 0/271 |
| 15-44yr | 167/263 (63.5) | 107/271 (39.5) |
| 45-64yr | 94/263 (35.7) | 150/271 (55.4) |
| ≥65yr | 2/263 (0.8) | 14/271 (5.2) |
| Female sex — no./total no. (%) | 129/263 (49.0) | 137/271 (50.6) |
| Occupation |  |  |
| Employee | 130/263 (49.4) | 92/233 (39.5) |
| Self-employed | 51/263 (19.4) | 39/233 (16.7) |
| Unemployed | 41/263 (15.6) | 44/233 (18.9) |
| Retired | 21/263 (8.0) | 23/233 (9.9) |
| Agricultural worker | 1/263 (0.4) | 25/233 (10.7) |
| Student | 11/263 (4.2) | 2/233 (0.9) |
| Medical staff | 8/263 (3.0) | 8/233 (3.4) |
| Smoking history — no./total no. (%) |  |  |
| Never smoked | 226/263 (85.9) | 252/271 (93.0) |
| Former smoker | 23/263 (8.7) | 8/271 (3.0) |
| Current smoker | 14/263 (5.3) | 11/271 (4.1) |
| Exposure history within past 14 days — no./total no. (%) |  |  |
| Huanan Seafood Wholesale Market | 4/263 (1.5) | 1/271 (0.4) |
| Contact with person with fever | 105/263 (40.0) | 54/271 (19.9) |
| Contact with COVID-19 patient in family or community | 110/263 (41.8) | 93/271 (34.3) |
| Chronic medical illness — no./total no. (%) |  |  |
| Hypertension | 24/263 (9.1) | 46/271 (17.0) |
| Diabetes mellitus | 9/263 (3.4) | 29/271 (10.7) |
| Hyperlipidemia | 3/263 (1.1) | 3/271 (1.1) |
| Cardiovascular and cerebrovascular diseases | 2/263 (0.8) | 16/271 (5.9) |
| Respiratory system disease | 18/263 (6.8) | 19/271 (7.0) |
| Hematological system disease | 1/263 (0.4) | 2/271 (0.7) |
| Chronic kidney disease | 0/263 | 3/271 (1.1) |
| Chronic liver disease | 14/263 (5.3) | 11/271 (4.1) |
| Autoimmune disease | 4/263 (1.5) | 6/271 (2.2) |

The median age of patients in Mobile Cabin Hospital was 40 years, ranging from 16– 68 years. The majority (167, 63.5%) were aged 15–44 years, and 94 (35.7%) were aged 45–64 years. The inpatients in Tongji Hospital were older, with a median age of 50 years (19–65 years); more than half (55.4%) were aged 45–64 years. There was a similar number of men and women (134/129 and 134/137, respectively) in the two hospitals. Half (49.4%) of the patients in Mobile Cabin Hospital were employed, and eight (3.0%) infected cases were medical staff; 25 (10.7%) patients in Tongji Hospital were agricultural workers, and eight (3.4%) inpatients were medical staff. Among all the subjects in Mobile Cabin Hospital and Tongji Hospital, most had a contact history with a person with a fever (40% and 19.9%, respectively) or with a confirmed COVID-19 case in the family or community (41.8% and 34.3%, respectively). Since Mobile Cabin Hospital was established for patients with mild symptoms and the ability to provide self-care, less than half of these patients had underlying diseases, including hypertension (9.1%), respiratory system disease (6.8%), chronic liver disease (5.3%), and diabetes mellitus (3.4%). More severe COVID-19 patients were admitted to Tongji Hospital. Thus, nearly half (49.8%) of the inpatients at Tongji Hospital had chronic medical illness, including hypertension (17.0%), diabetes mellitus (10.7%), and respiratory system (7.0%), cardiovascular, and cerebrovascular diseases (5.9%).

### Clinical characteristics of COVID-19 patients in Mobile Cabin Hospital and Tongji Hospital

The clinical characteristics, including the common symptoms of COVID-19, the radiologic findings of chest CT, and the PCR results of SARS-CoV-2 detection in nasopharyngeal swabs are summarized in Table 2.

**Table 2.** Clinical characteristics of COVID-19 patients in Mobile Cabin Hospital and Hospital.

| **Clinical characteristics** | **Mobile Cabin Hospital**<br>**(N=263)** | **Hospital**<br>**(N=271)** |
| --- | --- | --- |
| Symptoms — no. (%) |  |  |
| Fever | 107 (40.7) | 220 (81.2) |
| Highest temperature |  |  |
| <37.3°C | 156 (59.3) | 51 (18.8) |
| 37.3-38°C | 42 (16.0) | 58 (21.4) |
| 38.1-39.0°C | 56 (21.3) | 124 (45.8) |
| >39.0°C | 9 (3.4) | 38 (14.0) |
| Cough | 128 (48.7) | 220 (81.2) |
| Sore throat | 55 (20.9) | 54 (19.9) |
| Fatigue | 75 (28.5) | 135 (49.8) |
| Poor appetite | 62 (23.6) | 84 (31.0) |
| Chest stuffiness | 73 (27.8) | 104 (38.4) |
| Shortness of breath | 38 (14.4) | 50 (18.5) |
| Dyspnea | 17 (6.5) | 103 (38.0) |
| Nasal discharge | 17 (6.5) | 8 (3.0) |
| Nasal congestion | 20 (7.6) | 7 (2.6) |
| Headache | 45 (17.1) | 76 (28.0) |
| Myalgia or arthralgia | 56 (21.3) | 99 (36.5) |
| Diarrhea | 51 (19.4) | 117 (43.2) |
| Abdominal pain | 6 (2.3) | 47 (17.3) |
| Radiologic findings — no./total no. (%) |  |  |
| Chest CT |  |  |
| Single lung | 70/220 (31.8) | 24/267 (9.0) |
| Bilateral lungs | 129/220 (58.6) | 242/267 (90.6) |
| Normal | 21/220 (9.5) | 1/267 (0.4) |
| Laboratory findings — no. (%) |  |  |
| SARS-CoV-2 in throat swab specimens by RT-PCR |  |  |
| Positive | 159 (60.5) | 183 (67.5) |
| Probable positive | 18 (6.8) | 11 (4.1) |
| Negative | 86 (32.7) | 77 (28.4) |

**Table 3.** Ocular characteristics of COVID-19 patients in Mobile Cabin Hospital and Hospital.

| Ocular characteristics | Mobile Cabin Hospital<br>(N=263) | Hospital<br>(N=271) |
| --- | --- | --- |
| Symptoms — no. (%) |  |  |
| Conjunctival congestion | 10 (3.8) | 15 (5.5) |
| Conjunctival secretion | 28 (10.6) | 24 (8.9) |
| Ophthalmalgia | 15 (5.7) | 7 (2.6) |
| Foreign body sensation | 50 (19.0) | 13 (4.8) |
| Photophobia | 14 (5.3) | 1 (0.4) |
| Blurred vision | 55 (20.9) | 13 (4.8) |
| Dry eye | 82 (31.2) | 30 (11.1) |
| Tearing | 35 (13.3) | 20 (7.4) |
| Itching | 17 (6.5) | 36 (13.3) |
| Eye drops medication within past 14 days — no. (%) |  |  |
| Ofloxacin | 10 (3.8) | 11 (4.1) |
| Tobramycin | 3 (1.1) | 1 (0.4) |
| Ganciclovir | 1 (0.4) | 3 (1.1) |
| Artificial tears | 7 (2.7) | 5 (1.8) |
| Wearing glass or goggle — no. (%) | 96 (36.5) | 55 (20.3) |
| Hand-eye contact — no. (%) |  |  |
| Never | 65 (24.7) | 137 (50.6) |
| Seldom | 175 (66.5) | 111 (41.0) |
| Often | 23 (8.7) | 23 (8.5) |
| Total time of short-distance reading in a day — no. (%) |  |  |
| <2h | 20 (7.6) | 96 (35.4) |
| 2h-4h | 31 (11.8) | 66 (24.4) |
| 4h-8h | 88 (33.5) | 82 (30.3) |
| 8h-12h | 89 (33.8) | 19 (7.0) |
| >12h | 35 (13.3) | 8 (3.0) |
| Previous eye diseases — no. (%) |  |  |
| Conjunctivitis | 20 (7.6) | 13 (4.8) |
| Keratitis | 11 (4.2) | 3 (1.1) |
| Xerophthalmia | 21 (8.0) | 3 (1.1) |
| Cataract | 2 (0.8) | 7 (2.6) |
| Glaucoma | 4 (1.5) | 0 (0.0) |
| Macular disease | 0 | 1 (0.4) |
| Diabetic retinopathy | 1 (0.4) | 4 (1.5) |
| Other retinal disease | 1 (0.4) | 2 (0.7) |
| Optic nerve diseases | 1 (0.4) | 0 |

The clinical symptoms of the patients in the two hospitals were similar, but the percentages of involvement were various. The most common symptoms were fever (40.7% and 81.2% in Mobile Cabin Hospital and Tongji Hospital, respectively), cough (48.7% and 81.2%, respectively), and fatigue (28.5% and 49.8%, respectively). Of the patients in Tongji Hospital, 59.8% had a temperature over 38.1° C, and 43.2% of them had diarrhea. Only 6.5% of the subjects in Mobile Cabin Hospital suffered from dyspnoea, while 38.0% of the inpatients in Tongji Hospital developed dyspnoea while hospitalized. With regard to radiologic findings of the lung, most patients had bilateral involvement (58.6% and 90.6% in Mobile Cabin Hospital and Tongji Hospital, respectively). No radiologic feature of pneumonia was found in 21 (9.5%) patients in Mobile Cabin Hospital. All patients underwent SARS-CoV-2 detection via nasopharyngeal swabs several times. Respectively, 60.5% and 67.5% of patients were confirmed with positive SARS-CoV-2 detection.

### Ocular characteristics of COVID-19 patients in Mobile Cabin Hospital and Tongji Hospital

The ocular characteristics, including ocular symptoms, recent use of eye drop medication, eye protections, and previous eye diseases are summarized and presented in Table 2.

In Mobile Cabin Hospital, many patients suffered from ocular discomforts including conjunctival congestion (3.8%), increased conjunctival secretion (10.6%), ophthalmalgia (5.7%), foreign body sensation (19.0%), and tearing (13.3%). Eighty-two (31.2%) patients complained about dry eye. To relieve ocular symptoms, local treatments of ofloxacin (3.8%), tobramycin (1.1%), ganciclovir (0.4%), and artificial tears (2.7%) were given to the patients. A total of 36.5% patients in Mobile Cabin Hospital wore glasses or goggles. Most of them spent a lot of time on short-distance reading, especially on smartphones. A total of 33.5% patients spent 4–8 hours per day, and 33.8% of patients spent 8–12 hours per day reading. A few people had a history of eye diseases, including conjunctivitis (7.6%), keratitis (4.2%), and xerophthalmia (8.0%).

Among 271 patients in Tongji Hospital, 15 (5.5%) had conjunctival congestion during hospitalization. In addition, a total of 8.9% had increased conjunctival secretion, 2.6% had ophthalmalgia, and 4.8% had foreign body sensation. According to the evaluation by ophthalmologists, 11 (4.1%) and one (0.4%) inpatients received eye drops of ofloxacin and tobramycin, respectively. Local treatment of ganciclovir was given to three (1.1%) patients. Similar to patients in Mobile Cabin Hospital, 56 (20.3%) inpatients wore glasses or goggles. More than half (50.6%) of patients never touched their eyes with their hands. A total of 35.4% inpatients spent less than two hours per day on short-distance reading. A minority of inpatients had previous eye diseases, including conjunctivitis (4.8%), keratitis (1.1%), xerophthalmia (1.1%), cataract (2.6%), macular disease (0.4%), diabetic retinopathy (1.5%), and other retinal disease (0.7%).

### Ocular profiles and clinical characteristics of patients with conjunctival congestion

Figure 1 summarizes the ocular symptoms, eye drop medications, eye protections, history of eye disease, and clinical characteristics of all twenty-five patients with conjunctival congestion.

**Figure 1.**
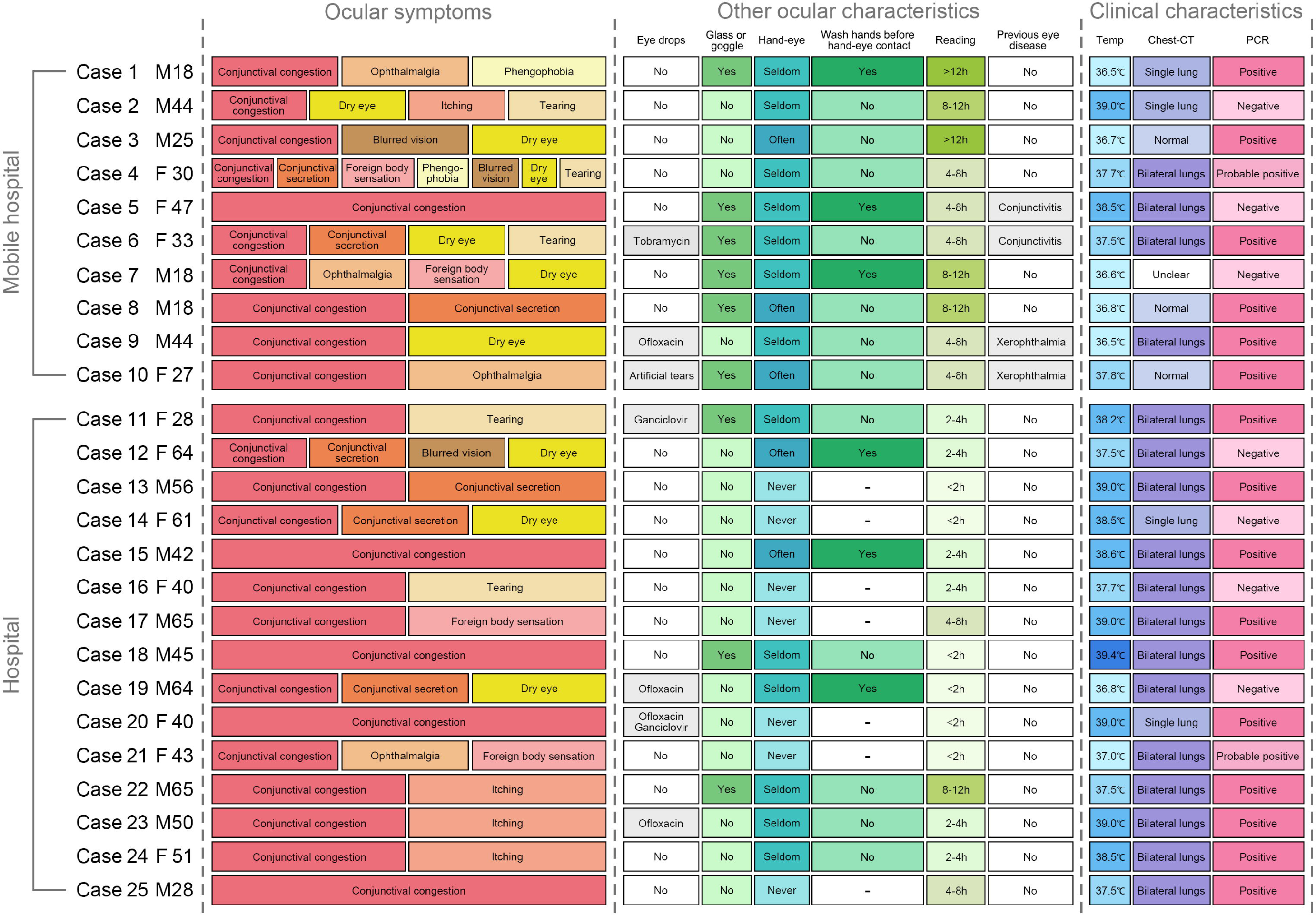
Detailed information about the ocular profiles and clinical characteristics of patients with conjunctival congestion. Twenty-five patients had conjunctival congestion. The detailed information includes ocular symptoms, eye protections, history of eye disease, and brief clinical characteristics.

Seven patients had conjunctival congestion and increased conjunctival secretion (cases 4, 6, 8, 12, 13, 14 and 19). Five patients had tearing (cases 2, 4, 6, 11 and 16). Four patients also had ophthalmalgia (cases 1, 7, 10 and 21), and four patients had foreign body sensation in the eyes (cases 4, 7, 17 and 21). Eighteen patients had touched their eyes with their hands, and twelve (cases 2, 3, 4, 6, 8, 9, 10, 11, 18, 22, 23 and 24) of them did not wash their hands before touching their eyes. Most of these patients had bilateral lung involvement indicated by chest CT (68%) and positive PCR results in SARS-CoV-2 detection (64%).

The duration of conjunctival congestion, onset date of clinical symptoms, and SARS-CoV-2 detection in patients with conjunctival congestion are summarized in Figure 2. The duration of conjunctival congestion ranged from two days to 12 days. Conjunctival congestion appeared mostly (72%) after the first clinical symptoms of COVID-19. Four patients (cases 11, 20, 23 and 24) had positive results for SARS-CoV-2 in nasopharyngeal swabs at the same time as the conjunctival congestion appeared. Three patients (cases 17, 21 and 25) had conjunctival congestion as an initial symptom.

**Figure 2.**
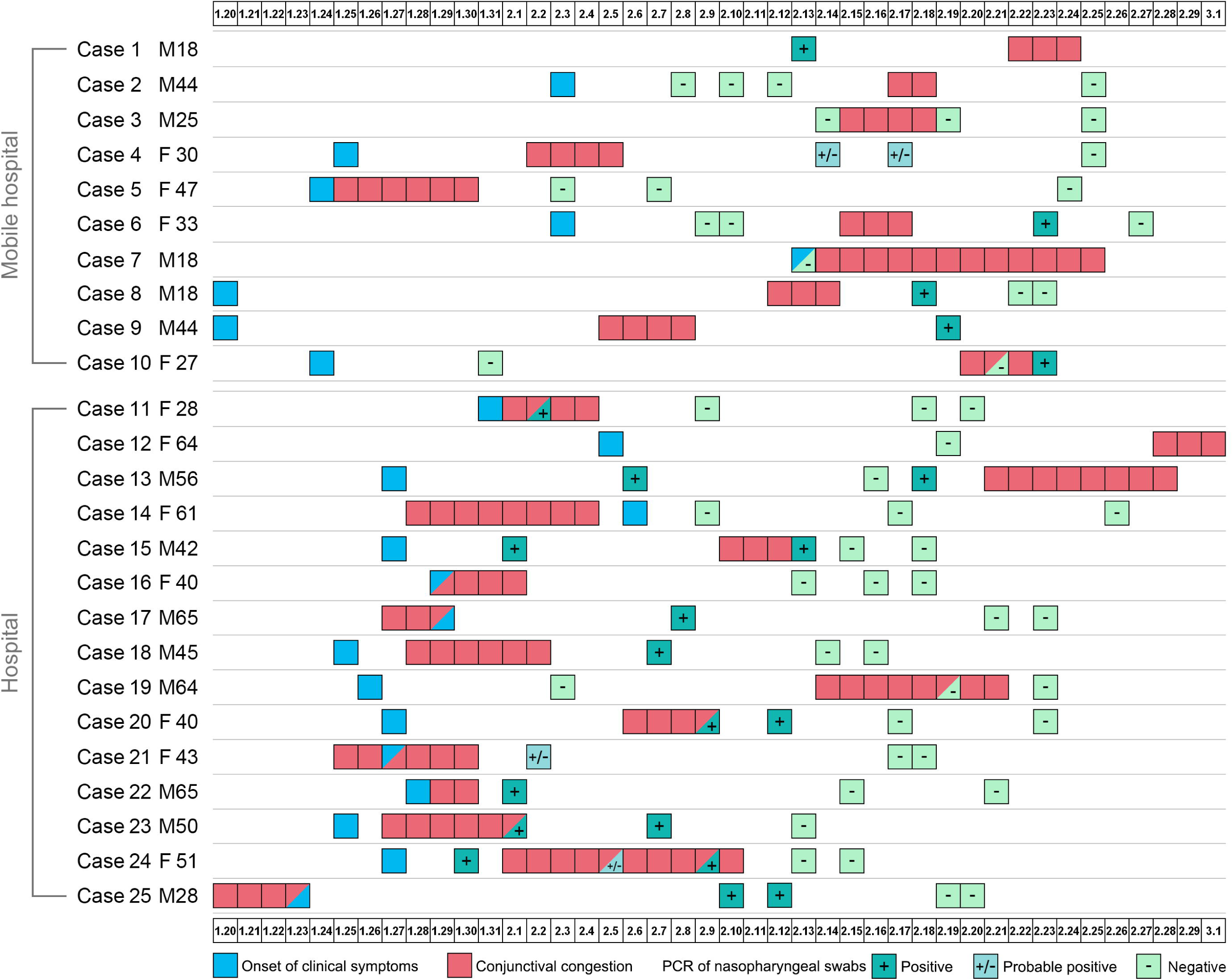
Detailed information about the duration of conjunctival congestion, the onset date of clinical symptoms, and SARS-CoV-2 detection in patients with conjunctival congestion. Twenty-five patients had conjunctival congestion. The numbers in boxes are calendar dates from January 20, 2019 to March 1, 2020. Blue box—onset date of the first clinical symptoms; red box—conjunctival congestion; green box**—**SARS-CoV-2 detection with different results.

## Discussion

To date, the epidemiologic data on the incidence of conjunctivitis in COVID-19 patients ranges from 0.82% to 4.76%.^1, 3, 9, 10^ However, the precise incidence of ocular manifestations relative to COVID-19 is unclear. In our present study, we found that 1) of a total of 534 COVID-19 patients identified, 25 patients (4.68%) presented with conjunctival congestion; 2) the incidence of dry eye (112, 20.97%), blurred vision (68, 12.73%), and foreign body sensation (63, 11.80%) ranked in the top three ocular symptoms; and 3) simultaneously, we also found that some COVID-19 patients had a history of eye diseases, among which the top five were conjunctivitis (33, 6.18%), dry eye (24, 4.49%), keratitis (14, 2.62%), cataract (9, 1.69%), and diabetic retinopathy (5, 0.94%).

Recent research has reported three cases of conjunctival congestion in 63 COVID-19 patients or suspected cases.^9^ Nine patients had conjunctival congestion among the 1,099 cases in Zhong and colleagues’ study.^1^ Consistent with their results, our previous single-center cross-sectional study showed that only two patients (2.78%) with conjunctivitis were identified from 72 patients with laboratory-confirmed COVID-19.^10^ However, these epidemiological investigations did not observe other ocular manifestations besides conjunctival congestion. Therefore, the present epidemiologic study summarized the manifestations present on the ocular surface. We enrolled 534 patients and found that COVID-19 patients exhibit ocular manifestations including conjunctival congestion (25), secretion (52), foreign body sensation (63), blurred vision (63), dry eye (112), itching (53), photophobia (15), and tearing (55). The incidence of conjunctival congestion in our study is 4.68%, which is higher than that in Zhong’s large sample report (9/1,099, 0.82%) and Xia’s study (1/30, 3.33%). Zhong and colleagues^1^ extracted data from 552 hospitals in 30 provinces, autonomous regions, and municipalities in China, while 21 common-type and nine severe-type COVID-19 patients were observed by Xia^3^ in Zhejiang province. However, the COVID-19 patients in our study mainly came from Tongji hospital (271) and Mobile Cabin Hospital (263) in Wuhan (the center of the SARS-CoV-2 outbreak), which may be the cause of the higher incidence of conjunctival congestion in the present study.

Given that SARS-CoV-2 nucleic acid was not detected in patients’ conjunctival swab sample, we did not diagnosis them with conjunctivitis directly. However, only four patients reported a history of eye disease among the 25 cases—two reported conjunctivitis and two reported dry eye. These 25 patients did not report any other eye disease history nor any symptoms associated with intraocular diseases (such as iritis, choroiditis, and retinal disease), which suggests that the possibility of endophthalmitis is very small, and conjunctivitis may be the primary cause of the conjunctival congestion. Moreover, conjunctival congestion and positive RT-PCR in pharyngeal swabs were found at the same time in four COVID-19 patients (cases 11, 20, 23, 24) who reported no eye disease history. Therefore, we suggest that the conjunctival swab test for SARS-CoV-2 should be performed in patients with conjunctival congestion. Our study also found that the average duration of conjunctival congestion was 4.9 ± 2.6 days (mean [SD]), ranging from 2 to 10 days. Two patients diagnosed with COVID patients had an initial symptom of conjunctival congestion, which reminds us that ocular manifestations occur early in the course of COVID-19. Therefore, healthcare workers should pay attention to patients’ ocular symptoms and manifestations in the early stage of disease and should perform a conjunctival swab test for SARS-CoV-2 in patients with conjunctival congestion.

SARS-CoV-2 is thought to be transmitted from person to person mainly through respiratory droplets or close contact.^11^ The ocular surface is exposed to the outside environment, which may become a potential gateway for pathogens such as viruses to invade the human body.^12, 13^ Thus, hand-eye contact should be avoided as much as possible. We found that a total of 332 COVID-19 patients had a history of hand-eye contact, including 286 cases who reported seldom hand-eye contact and 46 who reported frequent hand-eye contact. Among the 25 cases with conjunctival congestion, 18 (72%) had a history of hand-eye contact, 13 with frequent contact and 12 who never washed their hands, suggesting that hand-eye contact is possibly a high risk factor. Correlation analysis is warranted in future large-scale studies.

The present study also found that the incidence of dry eye, foreign body sensation, and blurred vision were the top three ocular symptoms in patients (20.9%, 11.8%, and 13.9%), which could be due to the fact that COVID-19 patients are more likely to have a lot of time to use electronic products. Our results showed that 321 of 534 COVID-19 patients (60.11%) spent more than four hours per day on short-distance reading. Time spent reading exceeded 12 hours per day in 43 patients (35 in Mobile Cabin Hospital and 8 in Tongji Hospital). Because of this, healthcare workers (particularly in Mobile Cabin Hospital) should propose that patients engage in some moderate physical exercise.

A portion of COVID-19 patients had a history of eye disease, among which the top five were conjunctivitis (6.18%), dry eye (4.49%), keratitis (2.62%), cataract (1.69%), and diabetic retinopathy (0.94%). As a result of the SARS-CoV-2 outbreak, it is difficult to provide rapid and effective treatment for concomitant eye disease in these COVID-19 patients. Therefore, we should attempt to take common eye disorders into consideration and pay more attention to these special patients to delay the progress of common eye diseases.

Consistent with previous studies, SARS-CoV-2–infected patients developed respiratory disorders with initial symptoms of fever, cough, chest stuffiness, and fatigue, which quickly progress to pneumonia and even shortness of breath.^8,14-16^ However, extra-pulmonary manifestations were also observed in a number of patients at the onset of the illness, including headache, myalgia or arthralgia, and diarrhea,^16^ and some even presented with asymptomatic infection.^17^ We also found that most of the enrolled COVID-19 patients had bilateral lung accumulation (371/524), and the SARS-CoV-2 RT-PCR test was positive at least once in 342 patients. However, the relationship between the ocular manifestations and the extra-ocular manifestations needs to be further studied.

The present study has some limitations. First, the sample size was small and the covered population consisted mainly of patients with mild disease; this is because only patients with mild symptoms and cured patients could complete our questionnaire survey. Second, this is a descriptive study, and no correlation analysis was performed; however, we present the ocular manifestations that we observed during the SARS-CoV-2 outbreak in detail. To date, this is the most comprehensive survey with the largest sample related to the eyes. Third, no normal population was observed in our study, therefore, a normal control group should be included for comparison in future studies.

In conclusion, although ophthalmology is not the main battlefield among all kinds of major infectious diseases, the significance of understanding the ocular manifestations of our present study lies in 1) helping to deepen the understanding of COVID-19–associated eye diseases; identify ocular symptoms, manifestations, and clinical outcomes; and enrich the symptom spectrum of COVID-19; 2) observing the incidence of eye diseases during COVID-19 treatment in hospital; and 3) providing a clue that patients with eye diseases are not being effectively treated during the COVID-19 outbreak. Our findings may provide useful information for the diagnosis and treatment of COVID-19 and indicate the need for effective eye protection for COVID-19 patients. Simultaneously, it also provides useful clues for the treatment of eye diseases in COVID-19 patients.

## Data Availability

Yes, we could provide all data referred to in the manuscriptif necessary.

## Acknowledgments

Hong Zhang, Xufang Sun designed and coordinated the study. Liwen Chen, Chaohua Deng and Xuhui Chen collected data. Liwen Chen performed and analyzed the data. Liwen Chen, Bo Chen, Xian Zhang prepared the figures and tables. Liwen Chen, Chaohua Deng and Xuhui Chen wrote the manuscript. All authors reviewed the results, revised the manuscript and approved it for submission. The authors declare that they have no conflicts of interest with the contents of this article.

